# Impact of Obesity on Catheter Ablation of Atrial fibrillation: Patient Characteristics, Procedural Complications, Outcomes and Quality of Life

**DOI:** 10.1101/2023.03.21.23287551

**Authors:** Chadi Tabaja, Arwa Younis, Pasquale Santageli, Medhat Farwati, Lorenzo Braghieri, Hiroshi Nakagawa, Walid I. Saliba, Ruth Madden, Patricia Bouscher, Mohamed Kanj, Thomas D. Callahan, David Martin, Mandeep Bhargava, Mina Chung, Bryan Baranowski, Shady Nakhla, Jakub Sroubek, Justin Lee, Tyler Taigen, Patrick J. Tchou, Oussama M. Wazni, Ayman A. Hussein

## Abstract

**Background:** Obesity is a well-known risk factor for atrial fibrillation (AF).

**Objective:** To evaluate the effect of baseline obesity on procedural complications, AF recurrence, and symptoms following catheter ablation (CA).

**Methods:** A total of 5841 patients undergoing AF ablation (2013-2021) were enrolled in a prospectively maintained registry. Primary endpoint was AF recurrence based on electrocardiographic documentation. Patients were categorized into 5 groups according to their baseline body mass index (BMI). Patients survey at baseline and at follow-up were used to calculate AF severity score (AFSS) as well as AF burden.

**Results:** Major procedural complications were low (1.5%) among BMI sub-groups. At 3 years AF recurrence was highest in Class III obesity patients (48%) followed by Class II (43%), whereas Class I, normal, and overweight had similar results with lower recurrence (35%). In multivariable analyses, Class III obesity was independently associated with increased risk for AF recurrence (HR=1.30, P=0.01), whereas other groups had similar risk in comparison to normal weight. Baseline AFSS was lowest in normal weight, and highest in Obesity-III, median [interquartile range] 10 [5-16] vs 15 [10-21]. In all groups, CA resulted in improvement in their AFSS with a similar magnitude among the groups. At follow-up, AF burden was minimal and did not differ significantly between the groups.

**Conclusion:** AF ablation is safe with a low complication rate across all BMI groups. Morbid obesity (BMI ≥40) was significantly associated with reduced AF ablation success. However, ablation resulted in improvement in QOL including reduction of the AFSS, and AF burden.

**What is known?:**

- Obesity is an independent risk factor of atrial fibrillation (AF)
- Catheter ablation (CA) has emerged as a standard of care in arrhythmia management, leading to improvements in quality of life, reductions in hospitalizations, and potential reductions in major adverse clinical outcomes.

More obese patients are being referred to catheter ablation of AF.

**What is new?:**

- AF ablation is safe with low complication rates across all BMI sub-groups (<1.5%)
- Morbid obesity (BMI ≥40) was associated with increased risk of AF recurrence after ablation
- Using patient reported outcomes, AF Ablation resulted in improvement of quality of life regardless of BMI

## Introduction

Obesity is a growing epidemic affecting nearly one third of the population worldwide.^1^ It is an independent risk factor for cardiovascular disease (CVD) and specifically atrial fibrillation (AF).^2-4^ Obesity is thought to promote AF through several mechanisms; most importantly via the inflammatory nature of epicardial adipose tissue, atrial remodeling and hemodynamic alterations that predispose to changes in cardiac morphology.^5^ In addition, obesity-related morbid conditions such as sleep apnea predispose to AF and its persistence.

Catheter ablation (CA) is currently widely accepted as the mainstay treatment for AF, with results showing improvement in quality of life (QoL), decreased hospitalizations, and potential improved cardiovascular outcomes.^6^ Along with the uptrend of obese patients being referred to CA, ^7^ it is important to explore the safety and efficacy of CA in such patients. Current studies pertaining to the impact of obesity on ablation outcomes are inconsistent. Some studies reported no impact ^8, 9^ while others reported reduced procedural success in obese patients.^10-12^ On the other hand, whether overweight or obese patients are more likely to develop procedure-related complications from CA is still debated. ^11, 13-15^ Furthermore, while it is widely accepted that the primary goal of AF ablation is to relieve symptoms, the impact of obesity on QoL improvement after CA is not well studied in the literature.

In this study, we aimed to investigate the safety, efficacy and outcomes among obese patients undergoing CA for AF in a prospectively maintained registry at a tertiary care center.

## Methods

### Patient population and data source

The study included patients from the Cleveland Clinic AF prospective registry who underwent AF CA between 2013 and 2021. All data, including but not limited to: clinical variables, procedural data, complications and assessment for arrhythmia recurrences, are prospectively collected in a maintained registry. In order to achieve individualized data, the current study included only patients undergoing a first time ablation at our institution. A total of 5841 consecutive patients with available data on body mass index (BMI) were included. The study was approved by the Cleveland Clinic Institutional Review Board.

### AF ablation protocol

Our approach to AF ablation was previously described.^16, 17^ In brief, antiarrhythmic medications were generally held before ablation and anticoagulants continued in an uninterrupted strategy peri-procedurally. The procedures were conducted under general anesthesia. Femoral venous access was obtained with ultrasound guidance. An intracardiac echocardiography (ICE) probe was positioned in the right atrium to help with catheter navigation, guide transseptal access to the left atrium (LA), and to monitor for procedural complications. Intravenous heparin was administered before LA transseptal access and continued during the procedure with a target activated clotting time (ACT) above 300 s. Three-dimensional electroanatomic maps were built to guide pulmonary veins isolation in addition to real-time monitoring of ablation catheter position and contact with ICE. Non-PV ablations were conducted at operators’ discretion. Concomitant atrial arrhythmia ablations were also performed as clinically indicated. Post-procedurally, patients were monitored for complications overnight and generally discharged within 24 h from procedure completion, unless longer stay was needed to further manage arrhythmias, procedural complications, or diuresis.

### Definitions, endpoints and post-ablation follow-up

Obesity was classified according to the BMI (body weight in kilograms divided by the square of body height in meters) and subdivided into categories as defined by the World Health Organization (WHO): the group of normal weight patients consisted of a BMI of 18.5–24.9 kg/m^2^ and the overweight group had a BMI of 25–29.9 kg/m^2^. Obesity was defined as Classes I through III (BMI: 30–34.9, 35–39.9, and ≥40 kg/m^2^), respectively with Class III obesity representing morbidly obese patients. Underweight patients constituted a very small percentage of total patients and hence were not included in the analysis.

The main outcome of interest was AF free survival following the blanking period (90 days from ablation). AF recurrence was defined in concordance with practice guidelines as electrocardiographic documentation of AF, lasting >30 s on 12-lead ECG, Holter recording, or during device interrogation of implanted devices. All clinical visits, correspondence with referring doctors, phone calls, and clinical letters were included in the follow-up data. Patients were provided with heart rhythm transmitters post-procedurally and instructed to transmit electrocardiographic recordings whenever symptomatic and on a weekly basis even when asymptomatic during the initial 3 months after ablation. Additional event recorder monitoring was performed beyond the initial 3-month period in patients with documented atrial arrhythmia or with symptoms suggestive of arrhythmia recurrence. Patients were also scheduled for follow-up visits with 12-lead ECG at 3, 6, and 12 months, and every 6 months thereafter. Additional visits were scheduled as needed for rhythm or anticoagulation management. Antiarrhythmics were used as necessary during the blanking period and generally discontinued whenever possible after the blanking period. Our comprehensive follow-up protocol has been described previously. ^16, 17^ In addition to arrhythmia monitoring and routine clinical assessments, patient-reported outcomes (PRO) were gathered at baseline and at each visit using an automated platform. For PRO assessment, we included a total of 2635 who participated in the PRO surveys at baseline and during follow up. The development of the PRO collection system was previously described in details ^18, 19^. This automated platform for PRO collection was developed and implemented at our institution to easily collect data following AF ablation with focus on QoL metrics of patients undergoing CA for AF. Patients automatically received email invitations to survey for AF-specific QoL outcomes within 10 days before the ablation procedures (baseline survey), 3 and 6 months after the procedure and every 6 months thereafter. The automated surveys included measurement of the AF Symptom Severity Scale (AFSS), a validated questionnaire for QoL assessment that quantifies symptom severity using 7 symptom-related questions. A score of 0 (no symptoms) to 5 (worse symptoms) is given to each question. Total scores range from 0 to 35; higher scores indicate worse symptoms and a greater negative impact on QoL. In addition to the AFSSS questionnaire, the automated surveys assessed AF burden (as indicated by AF frequency and duration). The AF frequency score ranged from 0 (no events) to 10 (continuous persistent AF). Similarly, the AF duration score ranged from 0 (no events) to 10 (continuous persistent AF).

All complication data were prospectively collected and adjudicated. Major procedural complications were defined in concordance with established guidelines as the occurrence of life-threatening complications and those requiring interventions or prolongation of hospital stay. These included the occurrence of cardiac tamponade, pericardial effusion requiring intervention, femoral pseudoaneurysm or arteriovenous fistula requiring intervention, bleeding requiring blood transfusion, hematoma requiring drainage, persistent phrenic nerve palsy, pulmonary vein stenosis (PVS), atrio-esophageal fistula, transient ischemic attack (TIA) or stroke, or death. Early readmissions or mortality within the first 4 weeks after ablation were also assessed.

### Statistical analyses

Continuous variables are expressed as mean ± SD. Categorical data are summarized as numbers (percent). Baseline clinical characteristics and complications were presented for each BMI group. The groups were compared with the use of the analysis of variance (ANOVA) or Wilcoxon ranked sum test for continuous variables; as appropriate, and χ^2^ test for categorical variables. Freedom from AF by different groups at baseline was displayed by tracing the Kaplan-Meier curves, with comparisons of cumulative event rates by the log-rank test.

To confirm associations determined by unadjusted analyses, adjusted Cox proportional hazards regression analysis was used to predict the associated risk in each BMI group when compared with normal weight group. The proportional hazards assumption was tested and met. Candidate covariates for the models are defined in **Supplementary Table**. Final model covariates were determined with the use of a stepwise selection process and were included if statistically significant at p <0.05 for the primary end point. The final proportional hazard regression models were adjusted for gender, age, AF type (paroxysmal or persistent), prior ablation, prior use of antiarrhythmic drugs (AADs), use of Cryoballoon and posterior wall isolation.

All statistical tests were 2 sided, and a P value of <0.05 was considered to be statistically significant. Analyses were carried out with the use of SAS software (version 9.4; SAS Institute, Cary, NC).

## RESULTS

### Baseline characteristics and procedural complications

A total of 5841 patients were included (17% normal weight, 34% overweight, 27% Class I, 13% Class II and 9% Class III). Their baseline characteristics are summarized in **Table 1**. The mean age of the overall patient population was 64 ±10 years and 66% were male. Obese patients were statistically younger than non-obese patients (64±9.8, 64±9.1 and 61.1±9.7 years for Class I through III vs 65±10 and 66±11 years in normal and overweight respectively, P<0.001). Atrial fibrillation was paroxysmal in 47% (PAF) and persistent in 53% (PersAF). An inverse relationship was observed between AF type and BMI. Normal weight patients were more likely to have paroxysmal AF (57%) whereas the rate in obesity Class III was significantly lower (38%, P<0.05). The prevalence of coronary and cerebrovascular disease risk factors such as hypertension, diabetes mellitus and heart failure were increasingly higher in overweight, obese and morbidly obese patients (P<0.001). The prevalence of atrial flutter decreased with increasing BMI (P<0.001). Valvular heart disease was present in 13.6% of patients at baseline.

**Table 1.**
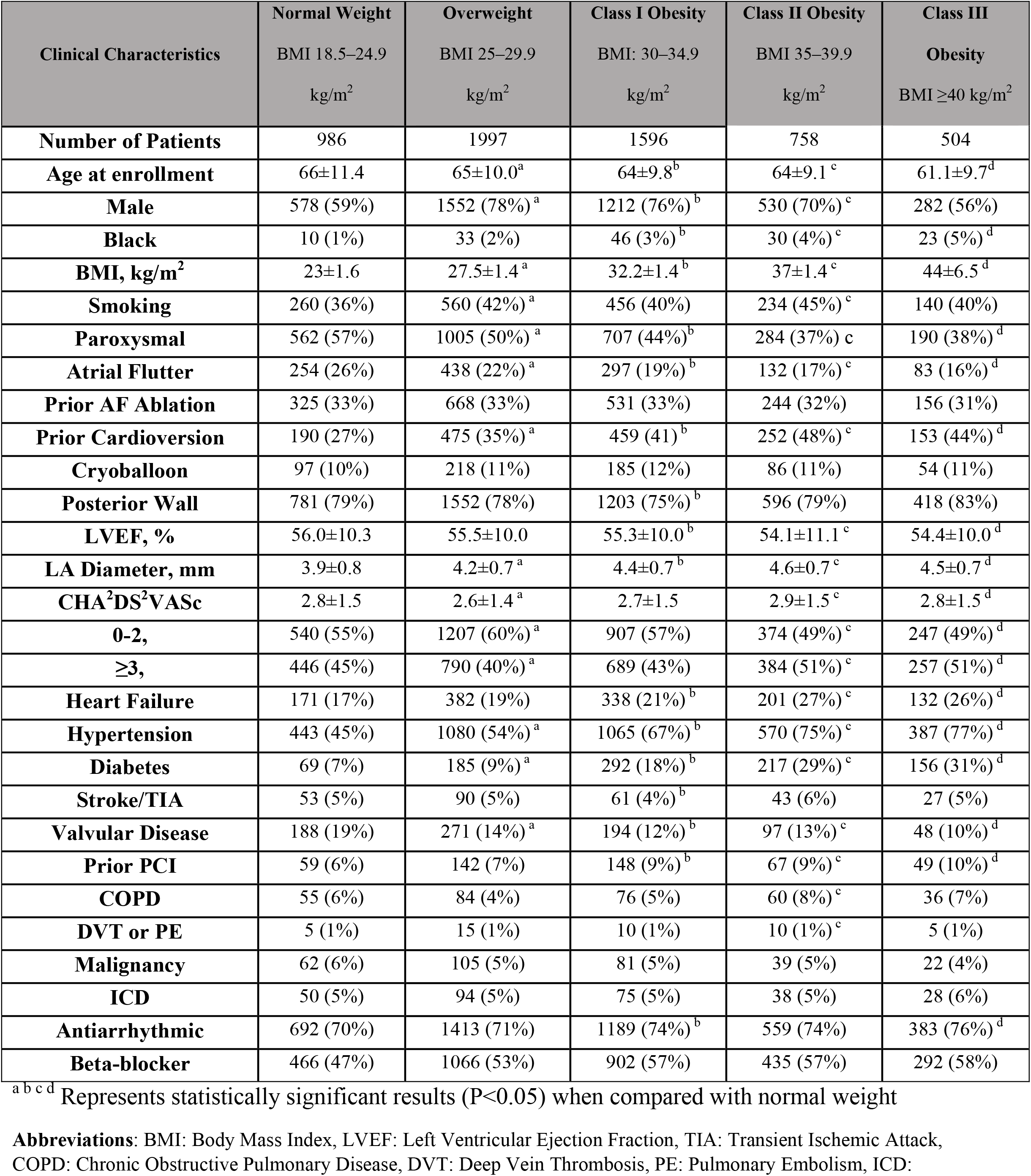
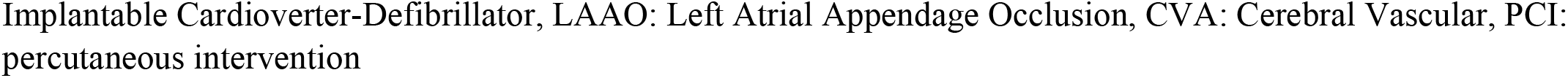
Clinical baseline characteristics according to body mass index categories.

| Clinical Characteristics | Normal Weight<br>BMI 18.5–24.9<br>kg/m<sup>2</sup> | Overweight<br>BMI 25–29.9<br>kg/m<sup>2</sup> | Class I Obesity<br>BMI: 30–34.9<br>kg/m<sup>2</sup> | Class II Obesity<br>BMI 35–39.9<br>kg/m<sup>2</sup> | Class III<br>Obesity<br>BMI ≥40 kg/m<sup>2</sup> |
| --- | --- | --- | --- | --- | --- |
| **Number of Patients** | 986 | 1997 | 1596 | 758 | 504 |
| **Age at enrollment** | 66±11.4 | 65±10.0<sup>a</sup> | 64±9.8<sup>b</sup> | 64±9.1<sup>c</sup> | 61.1±9.7<sup>d</sup> |
| **Male** | 578 (59%) | 1552 (78%)<sup>a</sup> | 1212 (76%)<sup>b</sup> | 530 (70%)<sup>c</sup> | 282 (56%) |
| **Black** | 10 (1%) | 33 (2%) | 46 (3%)<sup>b</sup> | 30 (4%)<sup>c</sup> | 23 (5%)<sup>d</sup> |
| **BMI, kg/m<sup>2</sup>** | 23±1.6 | 27.5±1.4<sup>a</sup> | 32.2±1.4<sup>b</sup> | 37±1.4<sup>c</sup> | 44±6.5<sup>d</sup> |
| **Smoking** | 260 (36%) | 560 (42%)<sup>a</sup> | 456 (40%) | 234 (45%)<sup>c</sup> | 140 (40%) |
| **Paroxysmal** | 562 (57%) | 1005 (50%)<sup>a</sup> | 707 (44%)<sup>b</sup> | 284 (37%)<sup>c</sup> | 190 (38%)<sup>d</sup> |
| **Atrial Flutter** | 254 (26%) | 438 (22%)<sup>a</sup> | 297 (19%)<sup>b</sup> | 132 (17%)<sup>c</sup> | 83 (16%)<sup>d</sup> |
| **Prior AF Ablation** | 325 (33%) | 668 (33%) | 531 (33%) | 244 (32%) | 156 (31%) |
| **Prior Cardioversion** | 190 (27%) | 475 (35%)<sup>a</sup> | 459 (41)<sup>b</sup> | 252 (48%)<sup>c</sup> | 153 (44%)<sup>d</sup> |
| **Cryoballoon** | 97 (10%) | 218 (11%) | 185 (12%) | 86 (11%) | 54 (11%) |
| **Posterior Wall** | 781 (79%) | 1552 (78%) | 1203 (75%)<sup>b</sup> | 596 (79%) | 418 (83%) |
| **LVEF, %** | 56.0±10.3 | 55.5±10.0 | 55.3±10.0<sup>b</sup> | 54.1±11.1<sup>c</sup> | 54.4±10.0<sup>d</sup> |
| **LA Diameter, mm** | 3.9±0.8 | 4.2±0.7<sup>a</sup> | 4.4±0.7<sup>b</sup> | 4.6±0.7<sup>c</sup> | 4.5±0.7<sup>d</sup> |
| **CHA<sup>2</sup>DS<sup>2</sup>VASc** | 2.8±1.5 | 2.6±1.4<sup>a</sup> | 2.7±1.5 | 2.9±1.5<sup>c</sup> | 2.8±1.5<sup>d</sup> |
| **0-2,** | 540 (55%) | 1207 (60%)<sup>a</sup> | 907 (57%) | 374 (49%)<sup>c</sup> | 247 (49%)<sup>d</sup> |
| **≥3,** | 446 (45%) | 790 (40%)<sup>a</sup> | 689 (43%) | 384 (51%)<sup>c</sup> | 257 (51%)<sup>d</sup> |
| **Heart Failure** | 171 (17%) | 382 (19%) | 338 (21%)<sup>b</sup> | 201 (27%)<sup>c</sup> | 132 (26%)<sup>d</sup> |
| **Hypertension** | 443 (45%) | 1080 (54%)<sup>a</sup> | 1065 (67%)<sup>b</sup> | 570 (75%)<sup>c</sup> | 387 (77%)<sup>d</sup> |
| **Diabetes** | 69 (7%) | 185 (9%)<sup>a</sup> | 292 (18%)<sup>b</sup> | 217 (29%)<sup>c</sup> | 156 (31%)<sup>d</sup> |
| **Stroke/TIA** | 53 (5%) | 90 (5%) | 61 (4%)<sup>b</sup> | 43 (6%) | 27 (5%) |
| **Valvular Disease** | 188 (19%) | 271 (14%)<sup>a</sup> | 194 (12%)<sup>b</sup> | 97 (13%)<sup>c</sup> | 48 (10%)<sup>d</sup> |
| **Prior PCI** | 59 (6%) | 142 (7%) | 148 (9%)<sup>b</sup> | 67 (9%)<sup>c</sup> | 49 (10%)<sup>d</sup> |
| **COPD** | 55 (6%) | 84 (4%) | 76 (5%) | 60 (8%)<sup>c</sup> | 36 (7%) |
| **DVT or PE** | 5 (1%) | 15 (1%) | 10 (1%) | 10 (1%)<sup>c</sup> | 5 (1%) |
| **Malignancy** | 62 (6%) | 105 (5%) | 81 (5%) | 39 (5%) | 22 (4%) |
| **ICD** | 50 (5%) | 94 (5%) | 75 (5%) | 38 (5%) | 28 (6%) |
| **Antiarrhythmic** | 692 (70%) | 1413 (71%) | 1189 (74%)<sup>b</sup> | 559 (74%) | 383 (76%)<sup>d</sup> |
| **Beta-blocker** | 466 (47%) | 1066 (53%) | 902 (57%) | 435 (57%) | 292 (58%) |

<sup>a b c d</sup> Represents statistically significant results (P<0.05) when compared with normal weight
**Abbreviations:** BMI: Body Mass Index, LVEF: Left Ventricular Ejection Fraction, TIA: Transient Ischemic Attack, COPD: Chronic Obstructive Pulmonary Disease, DVT: Deep Vein Thrombosis, PE: Pulmonary Embolism, ICD:
Implantable Cardioverter-Defibrillator, LAAO: Left Atrial Appendage Occlusion, CVA: Cerebral Vascular, PCI: percutaneous intervention

The procedural complication rates were low (<1.5%) and were similar in all groups (p>0.05). A breakdown of procedure related complications in BMI subgroups is provided in **Table 2**.

**Table 2.** Major procedural complications according to BMI group.

| **Clinical Characteristics** | **Normal** | **Overweight** | **Class I Obesity** | **Class II Obesity** | **Class III Obesity** | **Total** |
| --- | --- | --- | --- | --- | --- | --- |
| **Number of Patients, #** | 986 | 1997 | 1596 | 758 | 504 | 5841 |
| **Vascular or Bleeding** | 3(0.3%) | 2(<0.1%) | 5(0.3%) | 3(0.4%) | 2(0.4%) | 15 |
| **Pulmonary Vein Stenosis** | 3(0.3%) | 12(0.6%) | 9(0.5%) | 2(0.3%) | 0 | 26 |
| **Phrenic Nerve Injury** | 1(0.1%) | 7(0.3%) | 3(0.2%) | 2(0.3%) | 3(0.6%) | 16 |
| **Esophageal Fistula** | 1(0.1%) | 0 | 0 | 2(0.3%) | 0 | 3 |
| **Pericardial Effusion** | 6(0.6%) | 5(0.2%) | 4(0.2%) | 3(0.4%) | 1(0.2%) | 19 |
| **CVA or TIA** | 2(0.2%) | 1(0.05%) | 0 | 2(0.3%) | 0 | 5 |
| **Death within 30 Days** | 1(0.1%) | 1(0.05%) | 0 | 1(0.1%) | 0 | 3 |
| **Total** | 17(1.7%) | 28(1.4%) | 21(1.3%) | 15(1.9%) | 6(1.1%) | 87(1.5%) |

**Abbreviations:** CVA: Cerebral Vascular Event, TIA: Transient Ischemic Attack.

### Arrhythmia ablation outcomes and QOL by BMI type

During a mean follow up of 15±9 months, a total of 1533 patients (26%) had at least one episode of AF following their blanking period. **Figure 1** shows the Kaplan-Meier curves for freedom from AF by BMI groups. Upon 3 years of follow-up, the rate of AF recurrence for all BMI groups <35 kg/m^2^ ranged between 35 and 37%. The rate of AF recurrence was significantly higher in patients with BMI ≥35kg/m^2^ (Class II: 43%, Class III: 48%, P<0.05).

**Figure 1.**
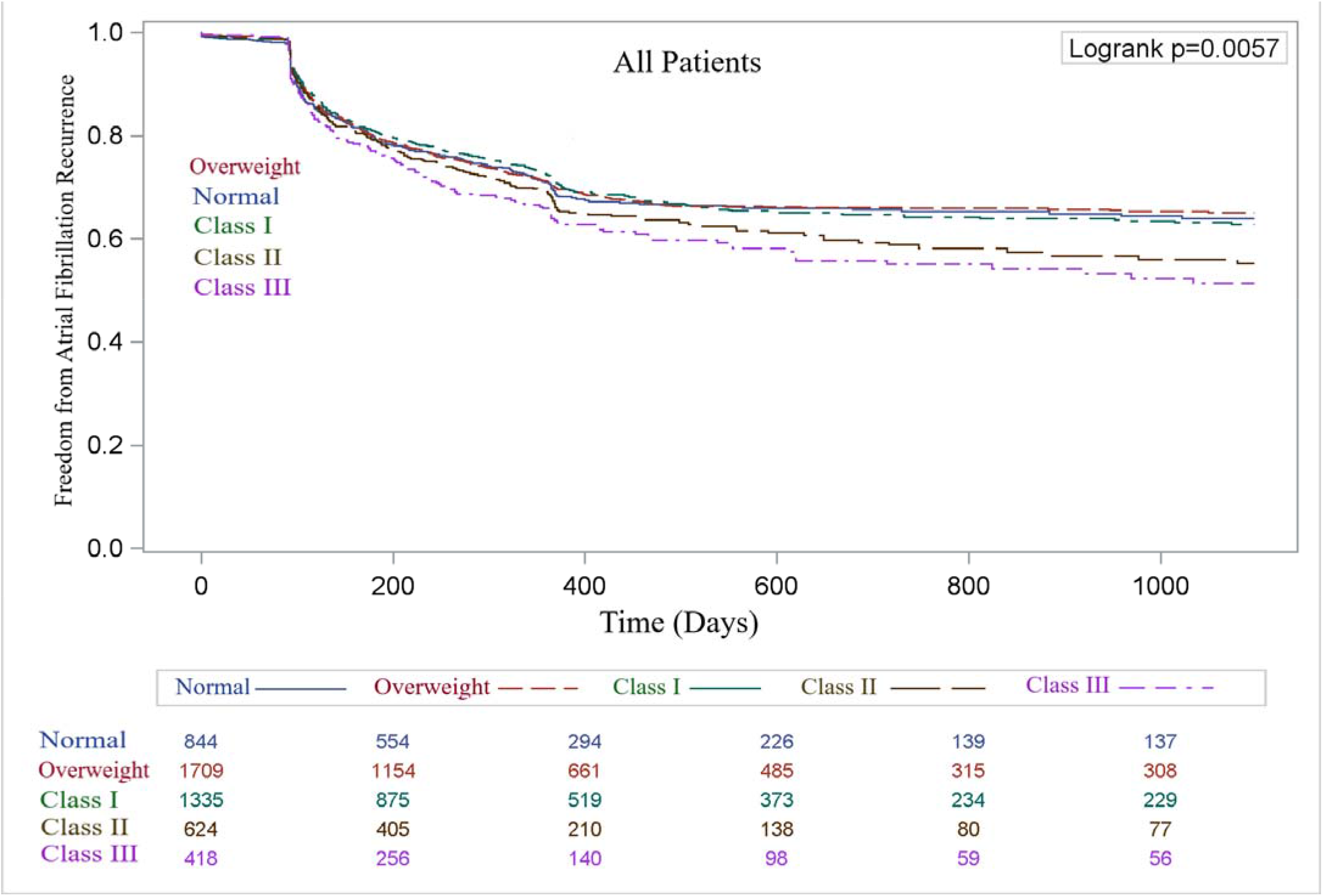
Kaplan–Meier curves showing rates of atrial fibrillation-free survival over time in normal weight (blue), overweight (red), and obesity classes: I (green), II (brown) and III (purple) subjects.

Unadjusted Cox proportional hazards analyses were performed to assess the association of different BMI groups with AF recurrence (normal BMI used as reference) (**Table 3)**. Before adjustment, only obesity Class III group (morbidly obese) was at increased risk of AF recurrence (HR= 1.30; 95% confidence interval [CI], 1.10-1.53, P=0.02) compared to normal BMI. In the adjusted model; which accounted for covariates that were found to have significant associations with AF recurrence in unadjusted analyses, Class III obesity was found to have an independent association with AF recurrence (HR=1.30, CI, 1.06-1.59, P=0.012). No significance difference was observed between the remaining BMI groups and those with normal BMI (overweight HR=0.98, Class I HR=0.98, Class II HR=1.16, P>0.05).

**Table 3.** Cox proportional hazards analyses assessing the association of BMI groups and recurrent arrhythmia after ablation.

| **Group** | **No. of Patients** | **No. of Events** | **HR** | **95% CI** | **p value** |
| --- | --- | --- | --- | --- | --- |
| **Unadjusted Analyses** |  |  |  |  |  |
| **Normal BMI** | 986 | 254 | Reference |  |  |
| **Overweight** | 1997 | 510 | 0.915 | 0.82-1.02 | 0.10 |
| **Class I Obesity** | 1596 | 398 | 0.94 | 0.83-1.05 | 0.25 |
| **Class II Obesity** | 758 | 213 | 1.14 | 0.99-1.32 | 0.07 |
| **Class III Obesity** | 504 | 155 | 1.30 | 1.10-1.53 | **0.002** |
| **Adjusted Analyses** |  |  |  |  |  |
| **All Patients** |  |  |  |  |  |
| **Normal BMI** |  |  | Reference |  |  |
| **Overweight** |  |  | 0.98 | 0.86-1.16 | 0.98 |
| **Class I Obesity** |  |  | 0.98 | 0.84-1.15 | 0.84 |
| **Class II Obesity** |  |  | 1.16 | 0.96-1.40 | 0.12 |
| **Class III Obesity** |  |  | 1.30 | 1.06-1.59 | **0.01** |

### Quality of Life

A total of 2635 patients participated in the automated surveys. In terms of PRO, there was a positive correlation between BMI and AFSS such as baseline AFSS was lowest in normal weight and overweight groups (median [interquartile range] AFSS was 10 [5-16]), and increasingly higher with higher obesity class (highest in obesity Class III, median [interquartile range] AFSS was 15 [10-21]; P<0.001).Upon follow-up, CA resulted in significant reduction in AFSS among all groups (median [interquartile range] normal weight 10 [5-16] vs 2 [0-7]; overweight 10 [5-16] vs. 3 [0-7]; Class I obesity 12 [7-18] vs 3 [0-8] Class II obesity 13.5 [8-19] vs 4 [1-10] and Class III obesity 15 [10-21] vs 5 [2-10]; P<0.05 for each comparison) (**Figure2**).

**Figure 2:**
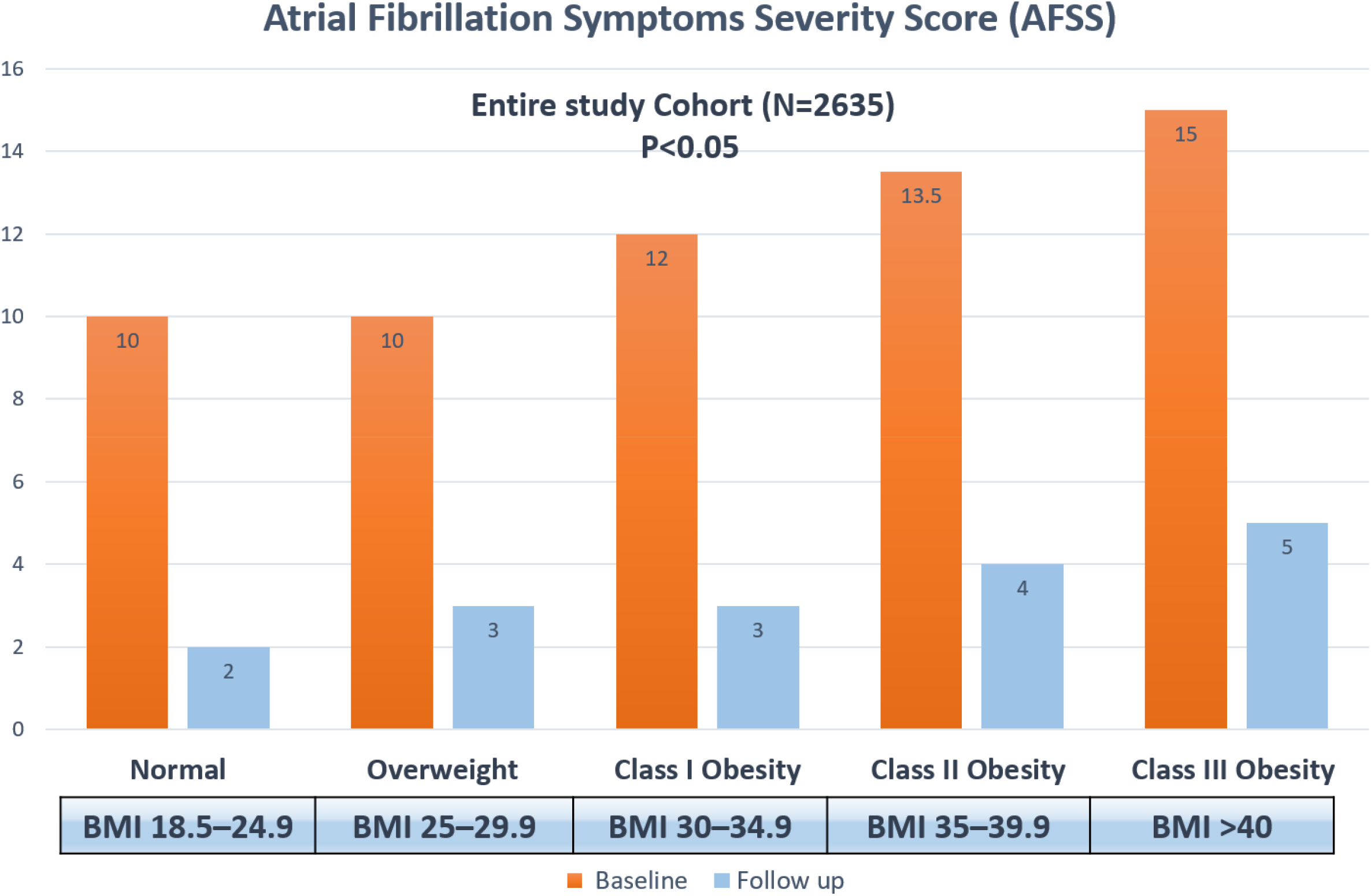
Atrial Fibrillation Symptom Severity Score according to BMI groups.

Similarly, AF burden scores were significantly reduced across all groups. The median AF duration score follow-up post-ablation was significantly lower than the median at baseline (median [interquartile range] normal weight 6.25 [5.62-8.75] vs 0 [0-2.5]; overweight 6.25 [6.25-10] vs. 0 [0-2.5]; Class I obesity 7.5 [7-10] vs 0 [0-2.5]; Class II obesity 8.75 [6.25-10] vs 0 [0-3.75] and Class III obesity 8.75 [6.25-10] vs 0 [0-3.75]; P<0.01 for each comparison). Similar results were obtained AF frequency score normal weight 7 [4-9] vs 0 [0-3]; overweight 7 [5-9] vs. 0 [0-2]; Class I obesity 8 [5-10] vs 0 [0-2]; Class II obesity 8 [5-10] vs 0 [0-2] and Class III obesity 8 [5-10] vs 0 [0-3]; P<0.05 for each comparison) **(Figure 3)**

**Figure 3:**
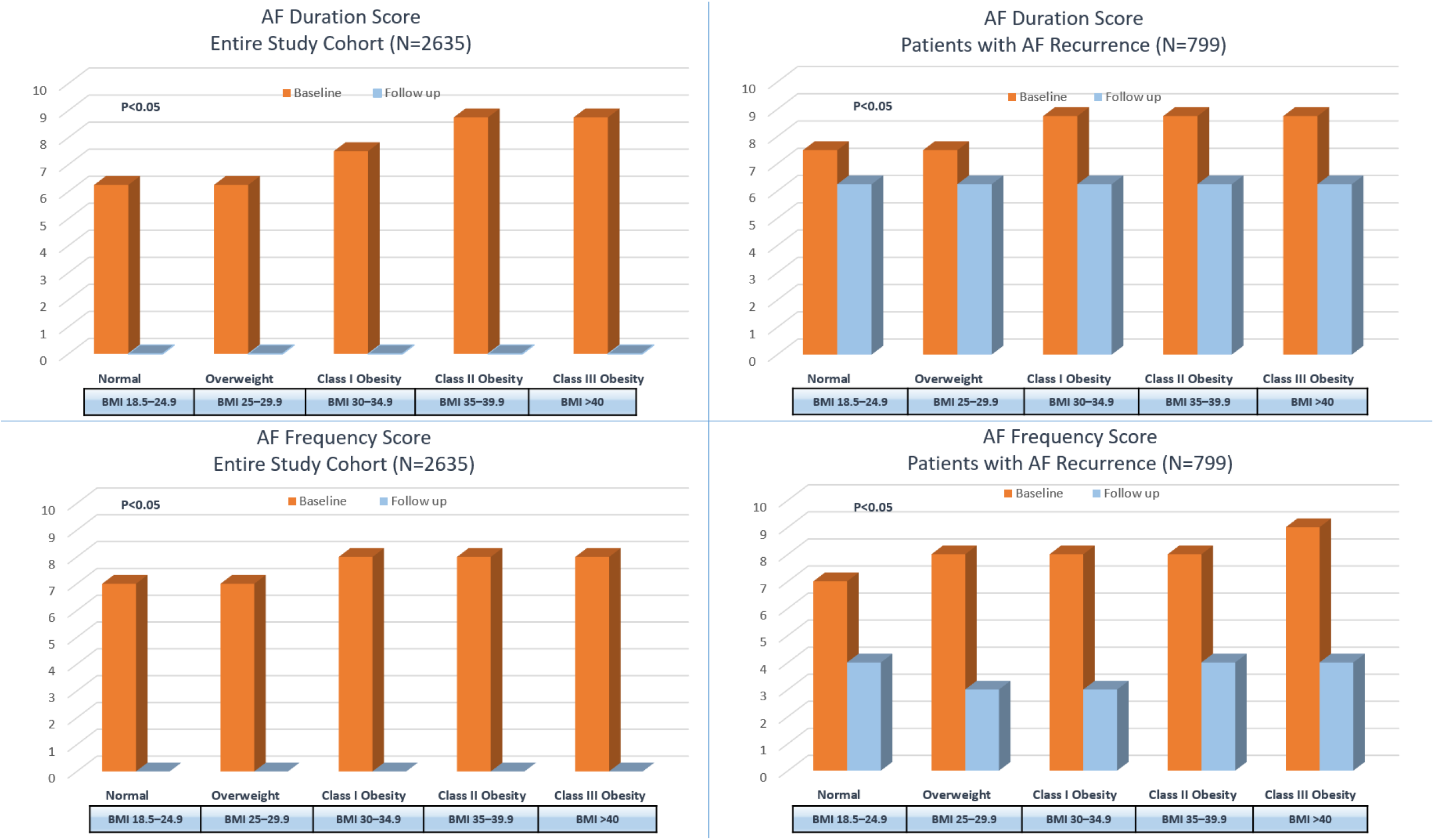
Atrial Fibrillation Duration and Frequency Scores in the entire study cohort and in those with arrhythmia recurrence.

In a subset, we further analyzed AF burden selectively in patients who had AF recurrence within during follow up (N=799) of CA and compared to baseline. Both AF duration score and AF frequency score were significantly reduced among patients with AF recurrence (median [interquartile range] AF duration score: normal weight 8 [4-10] vs 3 [2-7]; overweight 7.5[6.25-10] vs. 6.25 [2.5-8.75]; Class I obesity 8.75 [6.25-10] vs 6.25 [2.5-8.75]; Class II obesity 8.75 [6.25-10] vs 7.25 [6.25-10] and Class III obesity 8.75 [6.25-10] vs 6.25 [1.25-8.75]; median [interquartile range] AF frequency score: normal weight 7 [4-9.5] vs 3 [2-7]; overweight 8 [4-10] vs. 3 [2-7]; Class I obesity 8 [5-10] vs 3 [2-6]; Class II obesity 8 [5-10] vs 4 [2-8] and Class III obesity 9 [6-10] vs 4 [2-7.5]). (**Figure 3)**

## Discussion

In this prospective large cohort of patients undergoing CA for AF at a tertiary care center, we assessed the impact of obesity on procedural safety, outcomes and QOL. The main findings of the study are the following: 1) Patients with higher BMI have higher prevalence of comorbidities including hypertension, diabetes mellitus and heart failure and worse AFSS at baseline; 2) The safety profile of CA in the obese population is acceptable with similarly low complication rates among all BMI groups when compared to normal weight; 3) Class III obesity was independently associated with increased AF recurrence in adjusted analyses; 4) CA resulted in significant improvement in QoL and reduction in AF burden regardless of BMI group. To our knowledge this is the largest single-center cohort to investigate the impact of BMI on procedural outcomes, safety and QOL.

Catheter ablation has become standard of care in the management of AF, with excellent safety profiles; as demonstrated in this cohort from a tertiary care center. Importantly, our study showed that obesity did not increase the risk of AF ablation procedure-related complications. An important factor which could have contributed to this is obtaining vascular access with ultrasound in all patients ^20^ and the experience in the current report being from a high volume center. These data are in contrast with higher rates of complications reported from two large real-world nationwide inpatient sample studies ^21, 22^ which evaluated the impact of obesity on CA in more than 100,000 patients. Our data are in line with other studies which found no added procedure related risks in obese patients; ^8, 13, 14, 23^ even in morbidly obese which were previously reported to have higher complication rates. ^24^

In terms of clinical outcomes, published studies have produced conflicting findings on the association of obesity and arrhythmia recurrences after ablation. In the current report, only morbid obesity was independently associated with AF recurrence and in association with higher rates of persistent AF; potentially reflecting more advanced atrial myopathy, altered neuromodulation and hemodynamic strain. ^25^ Our results are consistent with prior reports, yet with minor nuances. Winkle et al. ^24^ divided 2715 AF patients undergoing ablation by BMI and reported a cut-off value of 35 kg/m^2^, beyond which BMI was found to have a negative impact on ablation outcomes. It is possible that such observation is primarily driven by BMI ≥ 40 kg/m^2^, as suggested by our findings. The same could apply to another study by Glover et al. ^26^ which studied the impact of BMI on AF ablation outcomes in 3333 patients, categorized into 3 groups (BMI < 25, BMI: 25.5–29, and BMI ≥ 30) and showed worse outcomes for patients with BMI≥30 kg/m^2^. Urbanek et al.^10^ investigated to role of cryoballoon ablation in obese patients with symptomatic AF and found that procedural success was reduced for patients with BMI>35 kg/m^2^. Another study by Sivasambu et al. ^27^ showed being overweight (BMI ≥ 25 to < 30) increased arrhythmia recurrence compared to normal weight patients. To be noted that the studies that did not find an association between obesity and ablation outcomes were limited by the fact that they combined all obese patients into a single category (>25 or >30) and included a smaller number of patients with short term follow-up.^8, 28^

While our findings indicate that ablation success is reduced with morbid obesity, it is unclear whether or not weight loss implementation as a pre-requisite before ablation would affect the outcomes. The recent SORT-AF ^29^ trial randomized patients with a BMI between 30 and 40 kg/m^2^ who underwent AF-ablation to usual care or weight reduction with a goal to evaluate the effect of weight loss on the success rate of AF ablation in obese patients. After ablation, AF burden did not differ between the two groups. Importantly, there was a significant decrease in AF burden among all BMI groups; in line with our study findings. Even though pre-procedural weight loss can have significant health benefits on morbidly obese patients undergoing ablation, it may be difficult to achieve in this group of patients who suffer from high AF burden and limited exercise capability. We suggest that AF ablation can still be performed in morbidly obese patients with the goal of reducing AF burden and improving QOL; while considering a formal weight management program and possibly bariatric surgery, as applicable.

## Limitations

The limitations of observational studies apply to the findings in this cohort, including the possibility of confounding from referral bias caused by patient selection. That being said, the study reflects real world clinical practice and data were collected prospectively. Another limitation pertains to asymptomatic AF recurrences which applies to most AF ablation studies. Nonetheless, AF ablation procedures are typically performed with the goal of improving QOL. The study showed a significant QOL benefit in all patients and regardless of BMI.

## Conclusion

In this cohort of AF patients undergoing ablation at a tertiary care center, complication rates were low and similar across BMI sub-groups. Morbid obesity with a BMI 40 kg/m^2^ was significantly associated with reduced AF ablation success. However, ablation resulted in improvement in QOL including reduction of the AFSS, and both duration and frequency of AF regardless of BMI. While it is reasonable to recommend AF ablation to symptomatic patients regardless of their BMI, the overall benefits of weight loss should not be overlooked.

## Data Availability

Data are available based on a reasonable request.

---

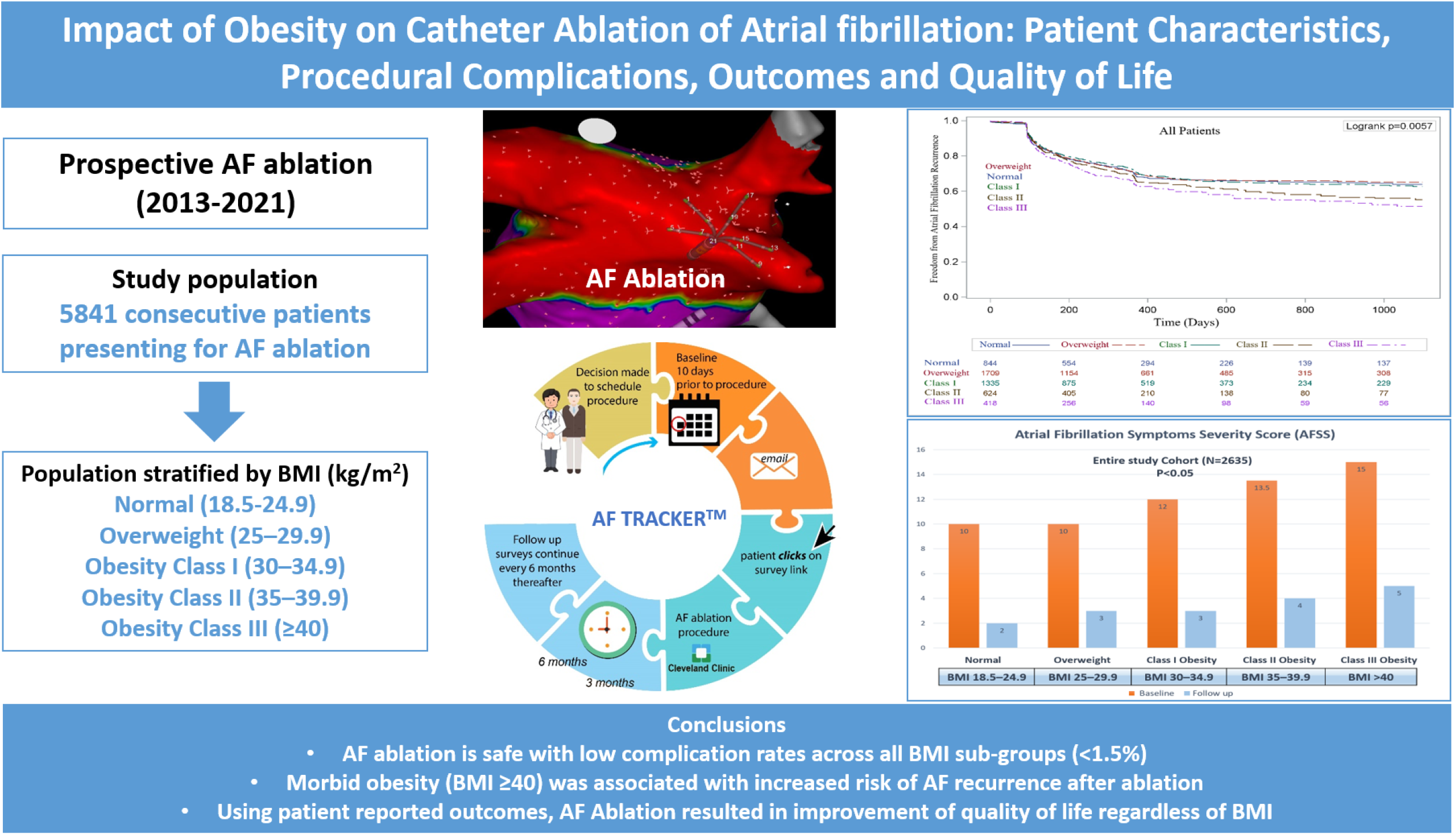

## Notes

### Competing Interest Statement

The authors have declared no competing interest.

### Funding Statement

This work has not been funded

### Author Declarations

Ourstudy was approved by the Cleveland Clinic Institutional Review Board.

